# Systematic and Statistical Review of Coronavirus Disease 19 Treatment Trials

**DOI:** 10.1101/2020.05.16.20102095

**Authors:** Juan A. Siordia, Michael Bernaba, Kenji Yoshino, Abid Ulhaque, Sooraj Kumar, Mario Bernaba, Edward Bergin

**Author notes:** Corresponding author. 329 N. Norris Ave., Tucson, Arizona 85719.

## Abstract

**Objective:** The following systematic review and meta-analysis compiles the current data regarding human controlled COVID-19 treatment trials.

**Methods:** An electronic search of the literature compiled studies pertaining to human controlled treatment trials with COVID-19. Medications assessed included lopinavir/ritonavir, arbidol, hydroxychloroquine, favipiravir, and heparin. Statistical analyzes were performed for common viral clearance endpoints whenever possible.

**Results:** Lopinavir/ritonavir showed no significant effect on viral clearance for COVID-19 cases (OR 0.95 [95% CI 0.50–1.83]). Hydroxychloroquine also showed no significant effect on COVID-19 viral clearance rates (OR 2.16 [95% CI 0.80–5.84]). Arbidol showed no seven-day (OR 1.63 [95% CI 0.76–3.50]) or 14-day viral (OR 5.37 [95% CI 0.35–83.30]) clearance difference compared to lopinavir/ritonavir. Review of literature showed no significant clinical improvement with lopinavir/ritonavir, arbidol, hydroxychloroquine, or remdesivir. Favipiravir showed quicker symptom improvement compared to lopinavir/ritonavir and arbidol. Heparin showed improvement with severe COVID-19 cases.

**Conclusion:** Current medications do not show significant effect on COVID-19 viral clearance rates. Favipiravir shows favorable results compared to other tested medications. Heparin shows benefit for severe cases of COVID-19.

## Introduction

Severe Acute Respiratory Syndrome – Coronavirus 2 (SARS-CoV2) is a novel coronarvirus responsible for causing Coronavirus Disease 19 (COVID-19). It quickly became a pandemic in the beginning of 2020. Originating in Wuhan, China, the virus rapidly spread to other countries of the world.[1] On January 30, 2020, the World Health Organization (WHO) declared SARS-CoV2 a Public Health Emergency of International Concern (PHEIC).[2] Medications are quickly being tested to assess for a suitable treatment regimen for the novel virus. The following systematic and statistical review assesses the current evidence regarding human controlled COVID-19 treatment trials.

## Methods

### Data Collection

An electronic search compiled human controlled studies analyzing treatments for COVID-19. Medical therapies investigated included lopinavir/ritonavir, arbidol, hydroxychloroquine, remdesivir, favipiravir, heparin, glucocorticoids, interferon, ivermectin, and convalescent plasma. Inclusion criteria included needing a control (whether standard therapy, placebo, or another medication) and testing among human subjects with COVID-19. In vitro and animal studies plus those without controls were not included in the review.

Databases included Google Scholar and Pubmed. Key words included: COVID-19, SARS-CoV2, randomized, controlled, human, retrospective, prospective, trial, chloroquine, hydroxychloroquine, lopinavir, ritonavir, arbidol, umifenovir, favipiravir, steroids, glucocorticoids, interferon, plasma transfusion, convalescent plasma, ivermectin, remdesivir, azithromycin, heparin, and low-molecular weight heparin. Abstracts and titles were reviewed for relevancy. Studies that had human subjects and COVID-19 Treatment a control arm were included in the study; otherwise, they were excluded. Duplicated studies were removed. The studies were organized based on the study medication; some studies presented more than one study medication and were included in more than one group. Statistical analysis was performed if there were two or more studies showing information regarding positive-to-negative conversion rates; number of days varied based on reported similarities among chosen studies.

### Statistical Analysis

If there were any common endpoints among the trials collected, a meta-analysis would then be performed. Endpoints were related to viral clearance. Statistical analyses used the Review Manager Version 5.3 (The Cochrane Collaboration, Copenhagen, Denmark) software program. A forest plot was created using the program with the DerSimonian and Laird fixed-effects model to reduce heterogeneity. The mean difference with a confidence interval (CI) of 95% was reported with the inverse variance method. Due to using a scale, the value marking no significance via confidence interval was zero. An I^2^ greater than 50% suggests significant heterogeneity. If there was significant heterogeneity, a random-effects model would be used instead.

## Results

### Study Selection

A total of 1780 articles were found with the keywords selected. A total of 24 studies were included initially based on title and abstract review. A total of 17 studies were included in the systematic review: Four studies elaborated about lopinavir/ritonavir; four studies studied arbidol, six for hydroxychloroquine, one for remdesivir, two for favipiravir, and two for heparin. Statistical analyses

COVID-19 Treatment regarding positive-to-negative conversion rates were possible for lopinavir/ritonavir, arbidol, and hydroxychloroquine. No human controlled trials were found for glucocorticoids, interferons, ivermectin, or convalescent plasma. Statistical analysis regarding positive-to-negative conversion rates was possible for lopinavir/ritonavir (two studies), arbidol (two studies), and hydroxychloroquine (four studies) (Figure 1).

**Figure 1:**
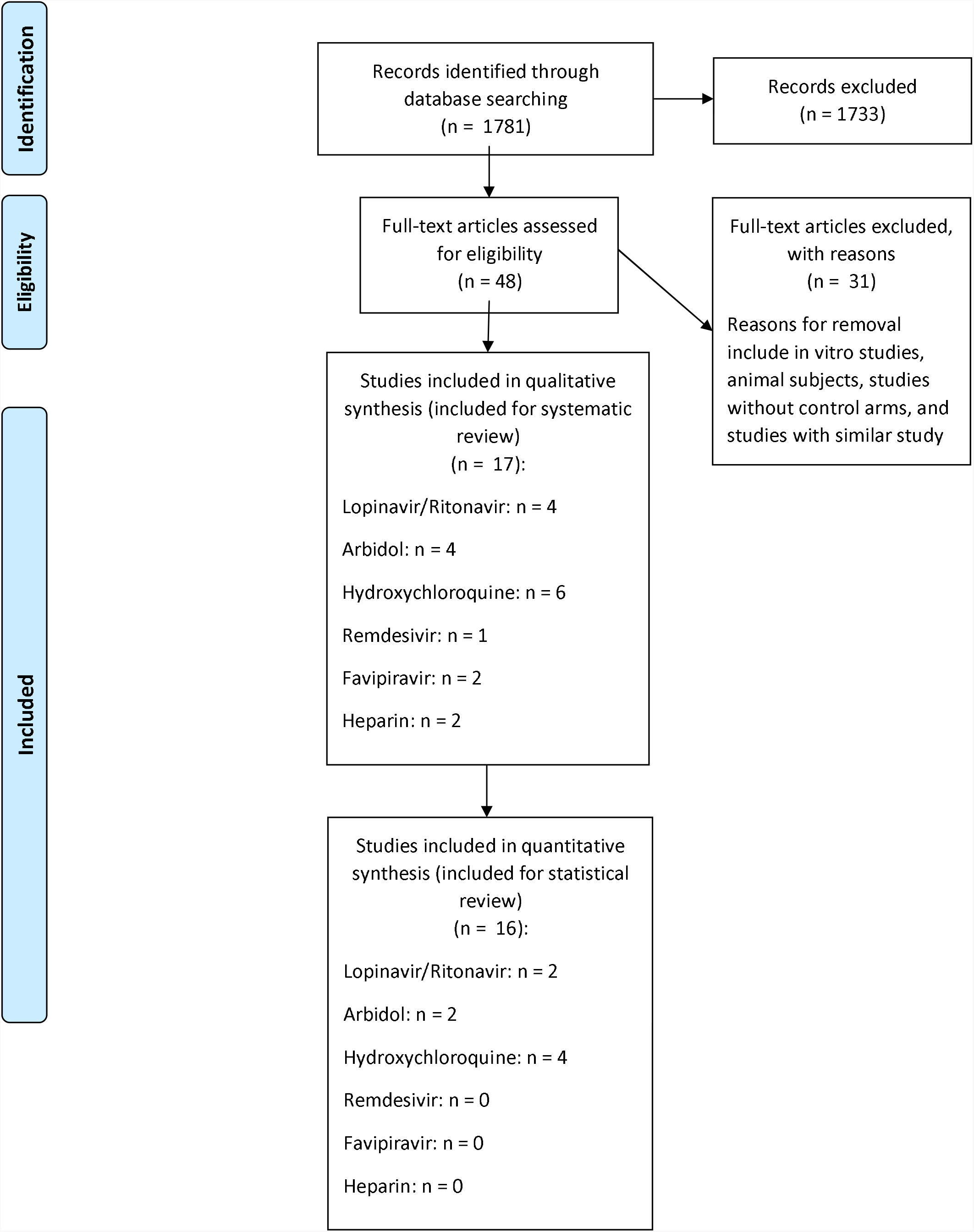
PRISMA Flowchart Revealing Study Selection Process

### Lopinavir/Ritonavir

#### Treatment

Four controlled trials exist regarding the treatment for COVID-19 (Table 1). Two studies are randomized controlled trials and two are retrospective controlled studies.[3–6]

**Table 1:**
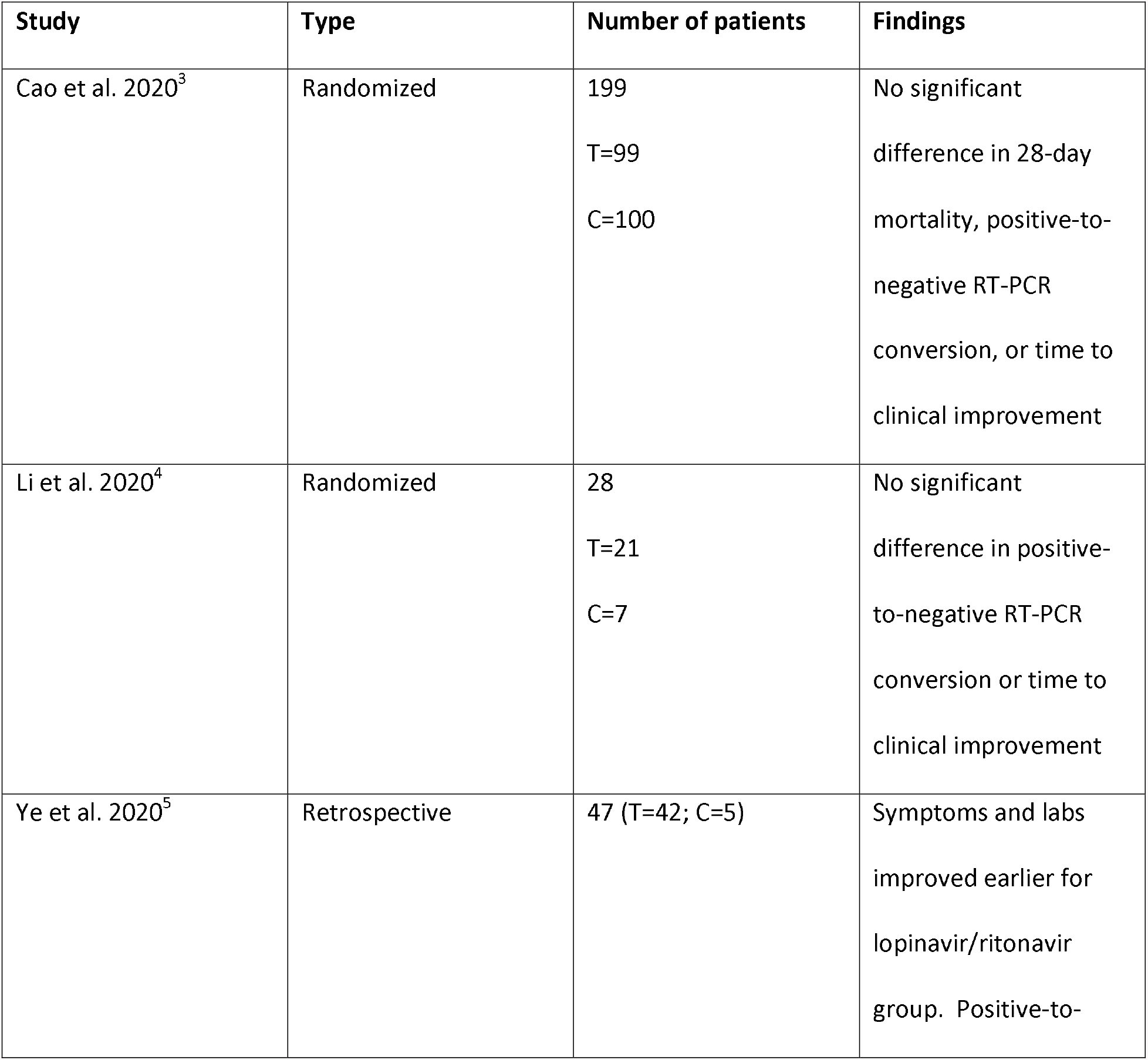
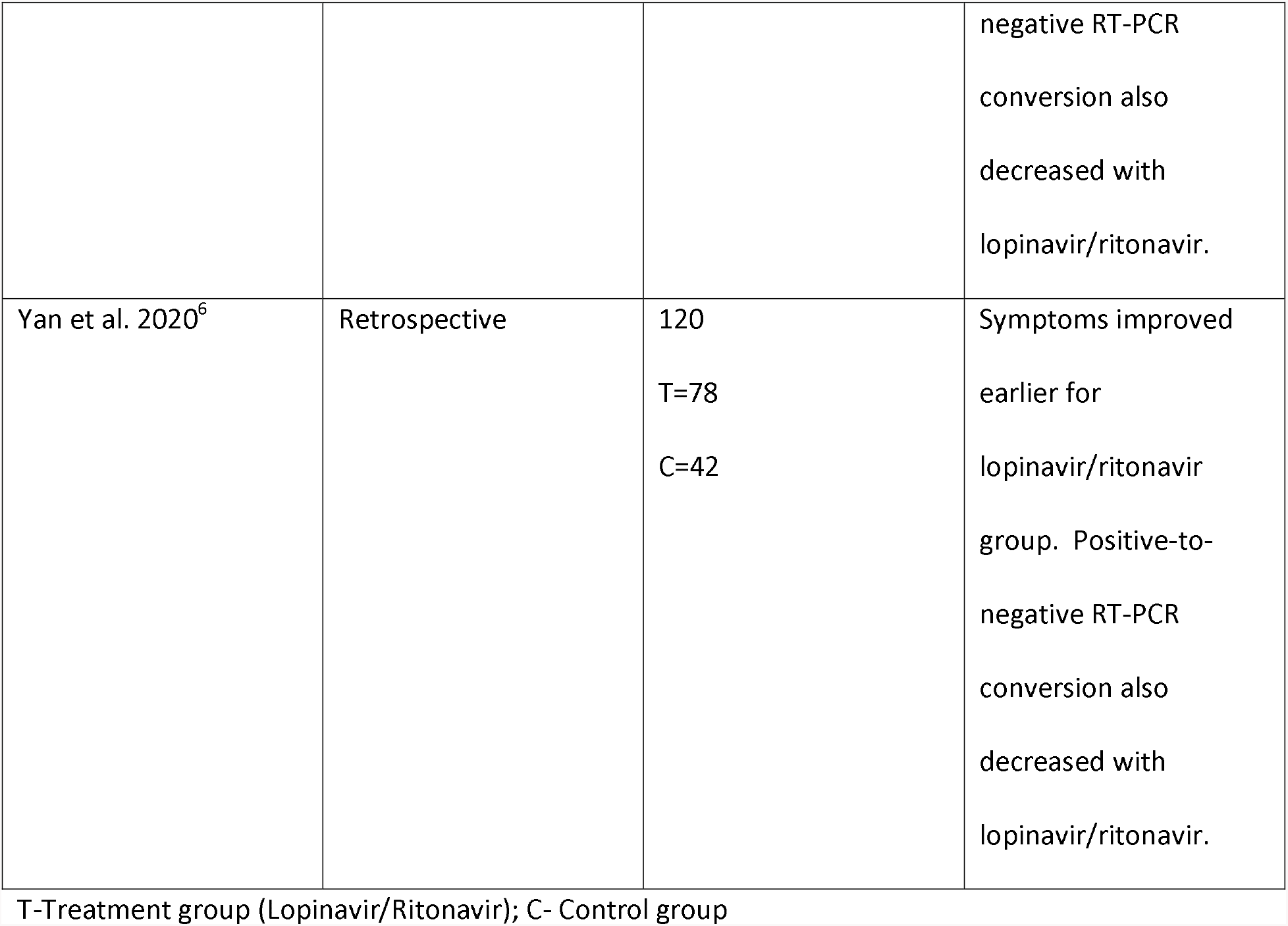
Characteristics of Lopinavir/Ritonavir Studies for COVID-19.

| Study | Type | Number of patients | Findings |
| --- | --- | --- | --- |
| Cao et al. 2020<sup>3</sup> | Randomized | 199<br>T=99<br>C=100 | No significant difference in 28-day mortality, positive-to-negative RT-PCR conversion, or time to clinical improvement |
| Li et al. 2020<sup>4</sup> | Randomized | 28<br>T=21<br>C=7 | No significant difference in positive-to-negative RT-PCR conversion or time to clinical improvement |
| Ye et al. 2020<sup>5</sup> | Retrospective | 47 (T=42; C=5) | Symptoms and labs improved earlier for lopinavir/ritonavir group. Positive-to- negative RT-PCR<br>conversion also<br>decreased with<br>lopinavir/ritonavir. |
| Yan et al. 2020<sup>6</sup> | Retrospective | 120<br>T=78<br>C=42 | Symptoms improved<br>earlier for<br>lopinavir/ritonavir<br>group. Positive-to-negative RT-PCR<br>conversion also<br>decreased with<br>lopinavir/ritonavir. |

T-Treatment group (Lopinavir/Ritonavir); C- Control group

The most recognized is the randomized, controlled, open-label trial by Cao et al.[3] The study showed no significant difference in terms of 28-day mortality or time of positive-to-negative reverse transcriptase-polymerase chain reaction (RT-PCR) conversion. Lopinavir-Ritonavir did reduce the time to clinical improvement by one day but was considered marginally non-statistically significant. This study had many limitations. The study was organized as an open label and with lack of placebo. About 14% of trial recipients could not complete a full 14-day treatment course due to adverse medication effects including nausea, vomiting, and diarrhea. However, the incidence of respiratory failure, acute kidney injury, and secondary infection was higher in the standard-care group.

Positive-to-negative RT-PCR conversion was not significant with lopinavir-ritonavir (Figure 2).[3–4] There was no significant difference between the study and control group at 14 days (OR 0.95 [95% CI 0.50–1.83]). Other retrospective studies suggest earlier clearance with lopinavir-ritonavir but did not report the results at 14 days.[5–6] Furthermore, the two studies that did suggest clearance are retrospective studies while the other two are randomized controlled trials.

**Figure 2:**
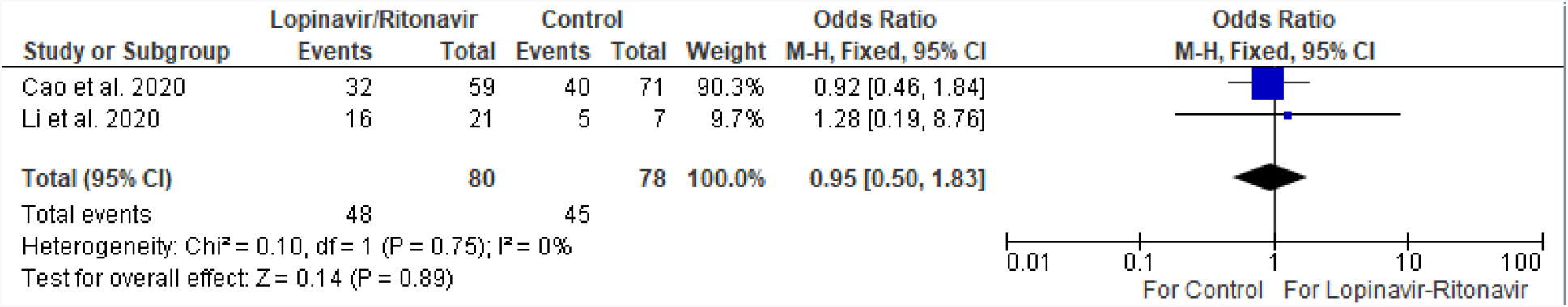
Positive-to-Negative RT-PCR Conversion of Lopinavir/Ritonavir versus Control at 14 Days.

#### Adverse Effects

The most significant lopinavir/ritonavir side effects include loss of appetite, nausea, vomiting, and diarrhea.[3–4] Diarrhea can possibly become severe.[4] Apart from elevated transaminase levels, other laboratory markers do not significantly differ from the control group.[3–4]

### Umifenovir (Arbidol)

#### Treatment

There are currently four controlled trials discussing the use of arbidol for the treatment of COVID-19 patients (Table 2).[4,7–9] Two of the trials are randomized while the other two are retrospective studies.

**Table 2:**
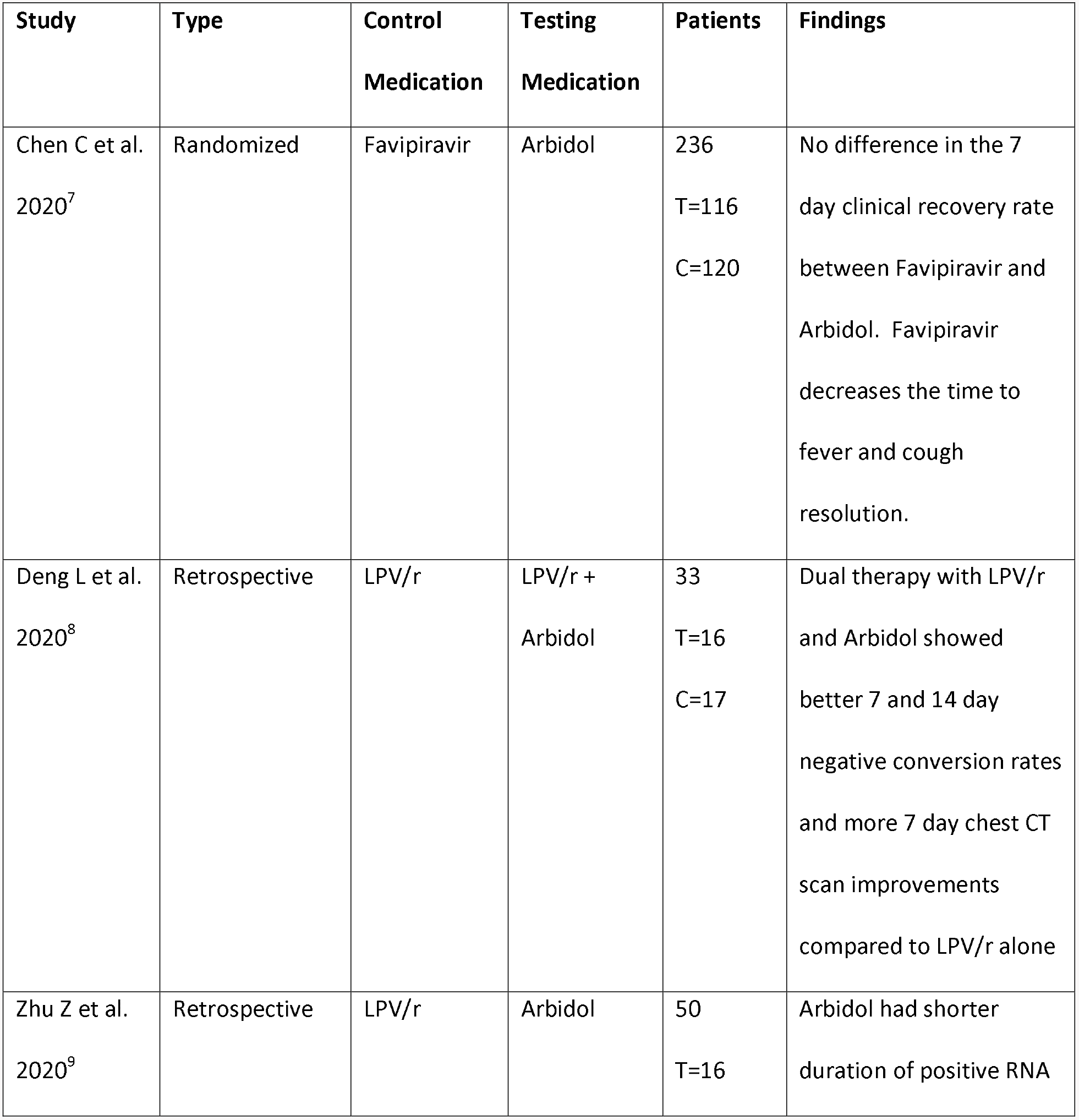
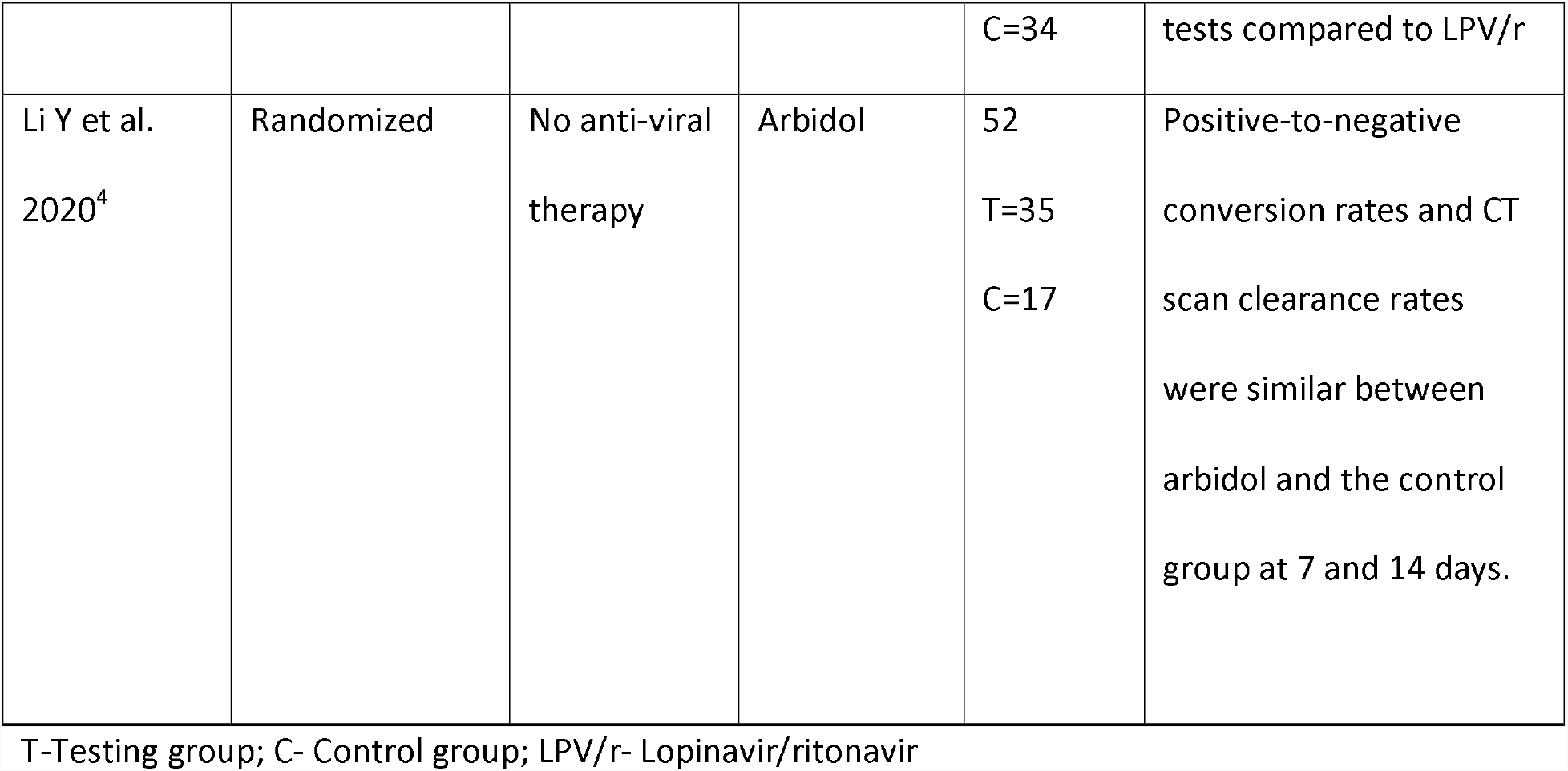
Characteristics of Arbidol Studies for COVID-19.

| **Study** | **Type** | **Control Medication** | **Testing Medication** | **Patients** | **Findings** |
| --- | --- | --- | --- | --- | --- |
| Chen C et al.<br>2020<sup>7</sup> | Randomized | Favipiravir | Arbidol | 236<br>T=116<br>C=120 | No difference in the 7 day clinical recovery rate between Favipiravir and Arbidol. Favipiravir decreases the time to fever and cough resolution. |
| Deng L et al.<br>2020<sup>8</sup> | Retrospective | LPV/r | LPV/r +<br>Arbidol | 33<br>T=16<br>C=17 | Dual therapy with LPV/r and Arbidol showed better 7 and 14 day negative conversion rates and more 7 day chest CT scan improvements compared to LPV/r alone |
| Zhu Z et al.<br>2020<sup>9</sup> | Retrospective | LPV/r | Arbidol | 50<br>T=16 | Arbidol had shorter duration of positive RNA |
|  |  |  |  | C=34 | tests compared to LPV/r |
| Li Y et al.<br>2020<sup>4</sup> | Randomized | No anti-viral<br>therapy | Arbidol | 52<br>T=35<br>C=17 | Positive-to-negative<br>conversion rates and CT<br>scan clearance rates<br>were similar between<br>arbidol and the control<br>group at 7 and 14 days. |

T-Testing group; C- Control group; LPV/r- Lopinavir/ritonavir

Only Li et al. includes a comparison between arbidol and standard supportive therapy.[4] The two retrospective studies include a comparison with lopinavir/ritonavir.[8–9] Chen et al. compare arbidol with favipiravir.[7]

Arbidol was commonly compared with lopinavir/ritonavir.[4,9] While there is no difference in positive-to-negative conversion rates between the two medications at the seventh (OR 1.63 [95% CI 0.76–3.50]) or 14^th^ day (OR 5.37 [95% CI 0.35–83.30]) (Figure 3 and Figure 4). Of note, for the 14-day comparison, a fixed effect model would show arbidol having more viral clearance compared to lopinavir/ritonavir (OR 5.0 [95% CI 1.50–16.64]). However, there was significant heterogeneity between the two studies (I^2^ = 66%). A random effects model was therefore employed to counter the heterogeneity, resulting in a nonsignificant difference between the two medications.

**Figure 3:**
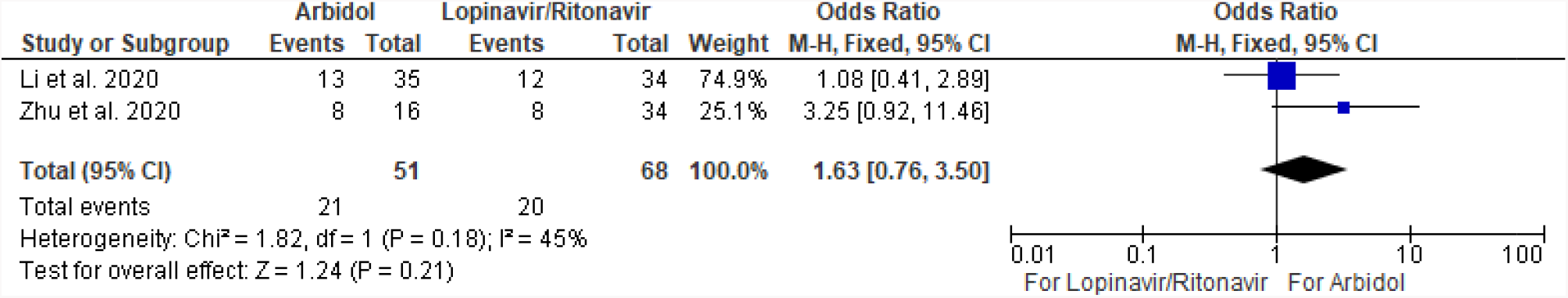
Positive-to-Negative RT-PCR Conversion of Arbidol versus Lopinavir/Ritonavir versus at 7 Days

**Figure 4:**
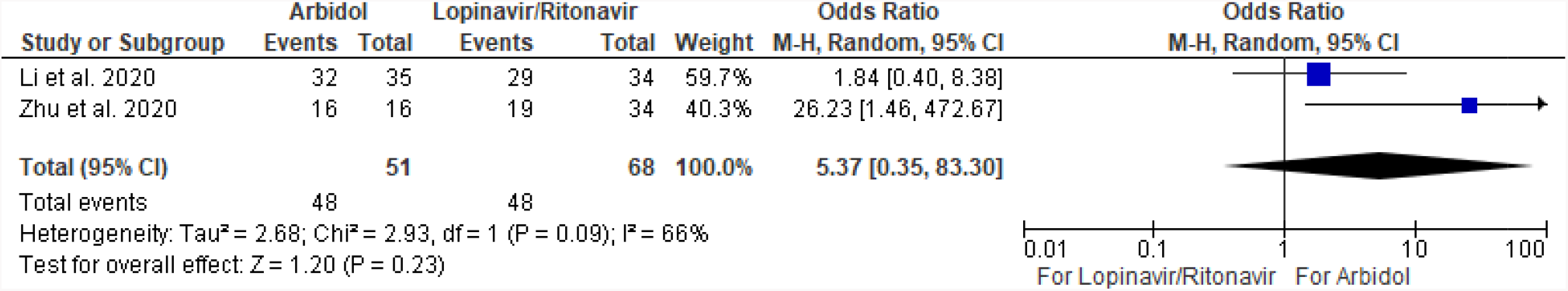
Positive-to-Negative RT-PCR Conversion of Arbidol versus Lopinavir/Ritonavir versus at 14 Days

Adding arbidol with lopinavir/ritonavir did show significant conversion rates and CT scan improvements compared to lopinavir/ritonavir by itself.[8]

While favipiravir did not show any difference compared to arbidol regarding seven-day recovery rate, it did show faster recovery from fever and cough. There was no difference regarding oxygen and noninvasive positive pressure ventilation use between arbidol and favipiravir.[7]

#### Adverse Effects

Arbidol side effects include nausea and diarrhea.[4] Arbidol demonstrated less hyperuricemia compared to favipiravir (p = 0.0014). Both favipiravir and arbidol did not show any significant difference in abnormal liver function tests, psychiatric symptom reaction, or digestive tract reactions.[7]

### Hydroxychloroquine

#### Treatment

Six controlled trials exist comparing hydroxychloroquine versus standard therapy (Table 3).[10–15] Three studies were randomized, one was prospective, and two were retrospective studies.

**Table 3:**
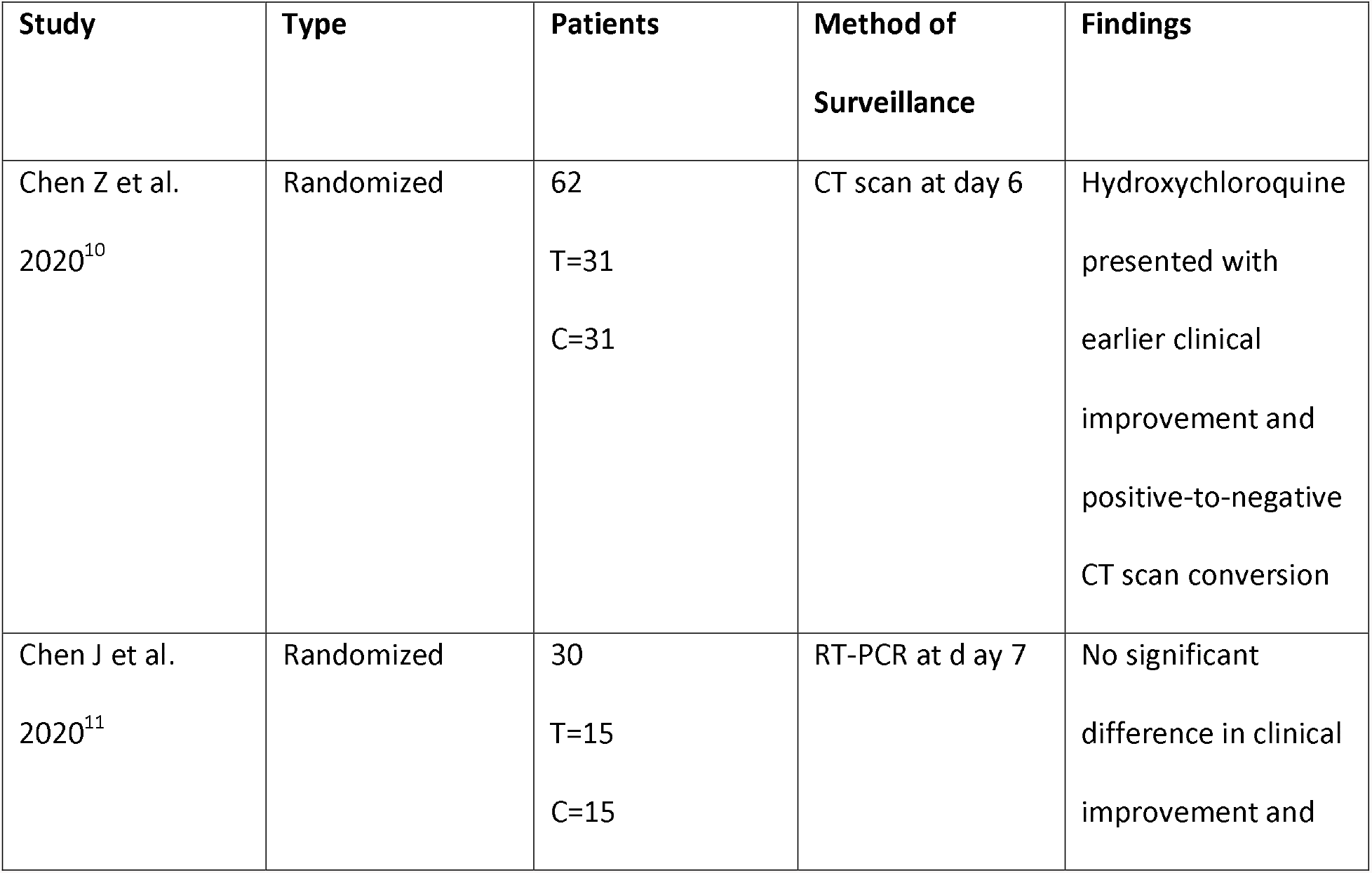
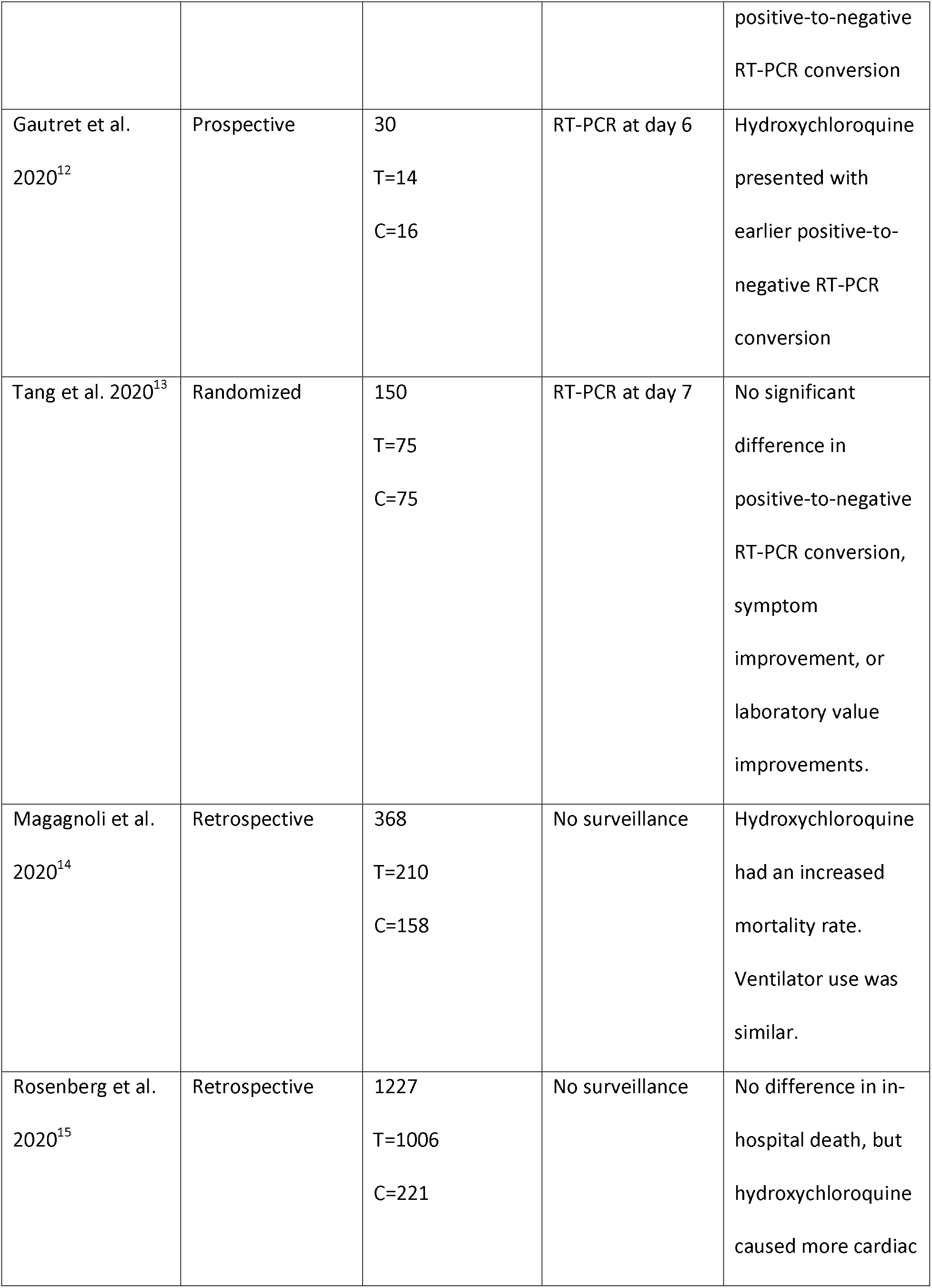
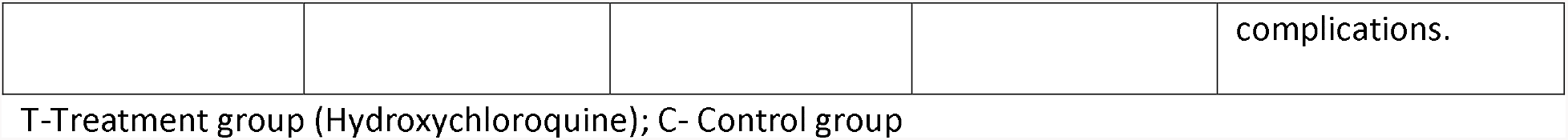
Characteristics of Hydroxychloroquine Studies for COVID-19.

| Study | Type | Patients | Method of Surveillance | Findings |
| --- | --- | --- | --- | --- |
| Chen Z et al.<br>2020<sup>10</sup> | Randomized | 62<br>T=31<br>C=31 | CT scan at day 6 | Hydroxychloroquine presented with earlier clinical improvement and positive-to-negative CT scan conversion |
| Chen J et al.<br>2020<sup>11</sup> | Randomized | 30<br>T=15<br>C=15 | RT-PCR at day 7 | No significant difference in clinical improvement and positive-to-negative<br>RT-PCR conversion |
| Gautret et al.<br>2020<sup>12</sup> | Prospective | 30<br>T=14<br>C=16 | RT-PCR at day 6 | Hydroxychloroquine<br>presented with<br>earlier positive-to-negative RT-PCR<br>conversion |
| Tang et al. 2020<sup>13</sup> | Randomized | 150<br>T=75<br>C=75 | RT-PCR at day 7 | No significant<br>difference in<br>positive-to-negative<br>RT-PCR conversion,<br>symptom<br>improvement, or<br>laboratory value<br>improvements. |
| Magagnoli et al.<br>2020<sup>14</sup> | Retrospective | 368<br>T=210<br>C=158 | No surveillance | Hydroxychloroquine<br>had an increased<br>mortality rate.<br>Ventilator use was<br>similar. |
| Rosenberg et al.<br>2020<sup>15</sup> | Retrospective | 1227<br>T=1006<br>C=221 | No surveillance | No difference in in-hospital death, but<br>hydroxychloroquine<br>caused more cardiac complications. |

T-Treatment group (Hydroxychloroquine); C- Control group

The data regarding hydroxychloroquine remains equivocal. The three randomized controlled trials present conflicting information regarding significance in clinical improvement and positive-to-negative conversion.[10–11,13] Chen Z et al. observed conversion based on CT scan results, but CT scans have a high negative predictive value for COVID-19 during the pandemic.[16–19] The prospective trial by Gautret et al. showed earlier conversion with hydroxychloroquine.[12] They included patients that took azithromycin with hydroxychloroquine in their study, but that was not included in this analysis. They have yet to present clinical status changes from their study.

A retrospective controlled study among veterans showed increased mortality with hydroxychloroquine use. Mechanical ventilation rates were similar among the two study arms.[14] Another retrospective review showed no difference in in-hospital mortality. [15]

The positive-to-negative conversion analysis (Figure 5) was performed at 6–7 days to include all the studies. RT-PCR or CT scans were used to monitor time to COVID-19 resolution. Hydroxychloroquine did not show significant effects on positive-to-negative conversion time compared to standard therapy (OR 2.16 [95% CI 0.80–5.84]). With significant heterogeneity (I^2^ = 56%), a random-effects model was used.

**Figure 5:**
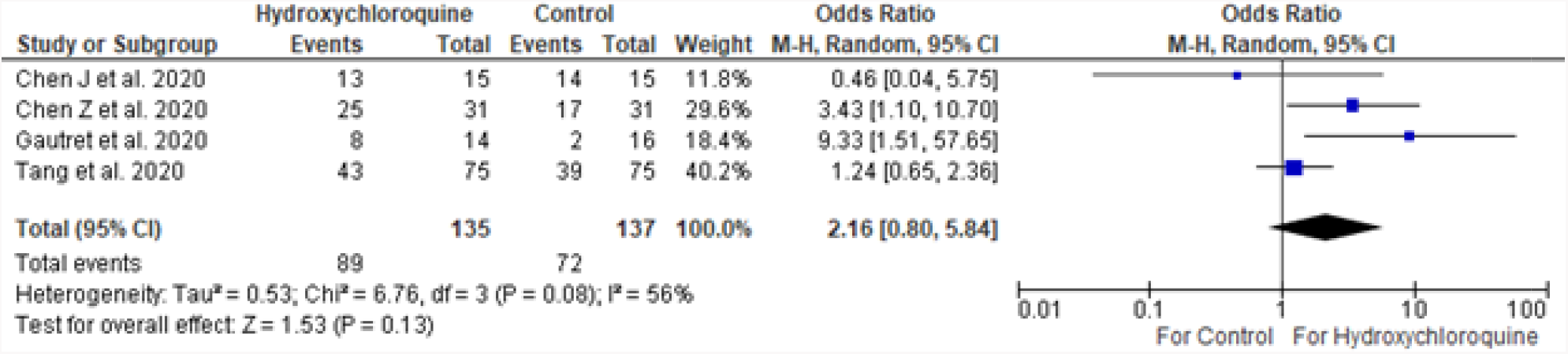
Positive-to-Negative Conversion of Hydroxychloroquine versus Control at 6–7 Days complications.

Analyzing the three randomized controlled trials only showed no significant difference between hydroxychloroquine and standard therapy (OR 1.50 [95% CI 0.88–2.57]) (Figure 6).[10–11,13] This was with nonsignificant heterogeneity (I^2^ = 38%), and therefore a fixed-effects model was kept.

**Figure 6:**
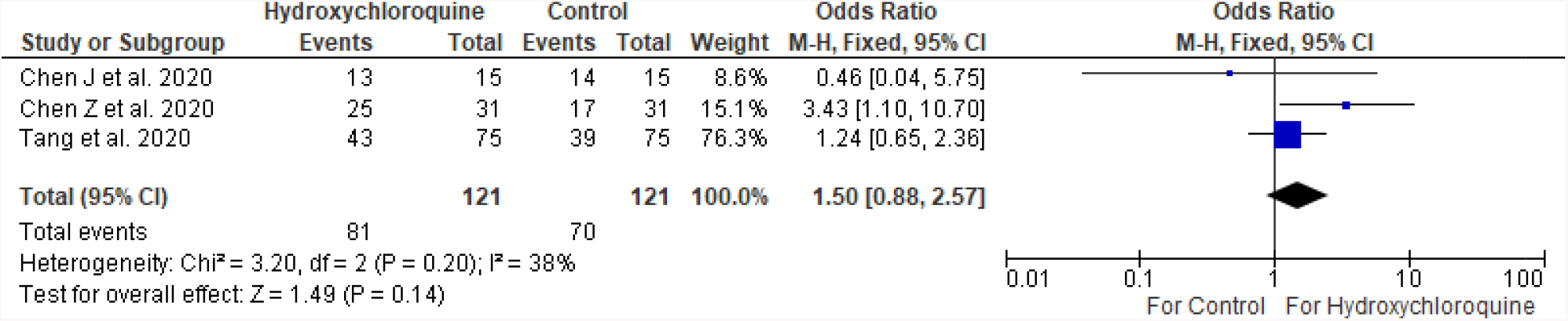
Randomized Controlled Trials showing Positive-to-Negative Conversion of Hydroxychloroquine versus Control at 6–7 Days

#### Adverse Effects

Cardiac complications, including cardiac arrest, were more common with hydroxychloroquine use especially when combined with azithromycin.[15] Gastrointestinal symptoms, including diarrhea and elevated transaminase levels, were mentioned with hydroxychloroquine, but they were not statistically significant compared to the control groups.[11–12,14]

### Remdesivir

#### Treatment

Currently there is only one published controlled trial with remdesivir (Table 4).[20] The randomized, double-blind, placebo-controlled trial showed no difference in time to clinical improvement compared to the control arm (Hazard ratio 1.23 [95% CI 0.87–1.75]).[20] A limitation of the study however was that patients in both groups were permitted concomitant use of lopinavir-ritonavir, interferons and/or corticosteroids.

**Table 4:** Characteristics of Remdesivir Studies for COVID-19.

| Study | Type | Patients | Findings |
| --- | --- | --- | --- |
| Wang et al. 2020<sup>20</sup> | Randomized | 237<br>T=158<br>C=79 | No difference with 28-day mortality, clinical improvement, and viral load change. |

T-Treatment group (Remdesivir); C- Control group

#### Adverse effects

About 66% who received remdesivir reported an adverse side effect. The most common side effects were constipation, hypoalbuminemia, hypokalemia, anemia, thrombocytopenia, and increased bilirubin.[20]

### Favipiravir

#### Treatment

There are two controlled trials regarding the use of favipiravir (Table 4).[7,21] The first is a randomized controlled trial comparing favipiravir to arbidol for COVID-19 patients.[7] Arbidol effects are similar to standard therapy.[4] The other is an open-label, non-randomized, prospective trial comparing favipiravir versus lopinavir/ritonavir.[21] Lopinavir/ritonavir is also similar to standard therapy.[3–4] Chen et al. showed no significant seven-day recovery rate with favipiravir compared to arbidol. The secondary endpoints of fever and cough relief did resolve significantly sooner with favipiravir compared to arbidol, with fever resolving for all patients at day 4 (versus day 7–8) and cough improving at day 8 (versus day 8+). There was no difference regarding oxygen and non-invasive positive pressure ventilation use.[7]

Cai et al. showed faster CT scan improvement and viral clearance with favipiravir compared to lopinavir/ritonavir. At day 14, 32/35 (91.43%) favipiravir subjects had improved chest CT scans compared to 28/45 (62.22%) lopinavir/ritonavir patients (p = 0.004). Viral clearance was sooner at 4 days with favipiravir compared to 11 days with lopinavir/ritonavir (p<0.001).[21]

#### Adverse Effects

Favipiravir shows a similar side-effect profile as to lopinavir/ritonavir, including nausea, vomiting, diarrhea, rash, and elevated transaminase levels.[7,21] Compared to arbidol, it increases uric acid levels more. While the side effect profile is similar to lopinavir/ritonavir, the frequency of adverse effects are less with favipiravir.[21]

### Unfractionated and Low-molecular weight heparin

#### Treatment

Two retrospective controlled studies included data regarding heparin use (Table 5).[22–23] These studies involved deep vein thrombosis prophylaxis dosing of unfractionated (15000 IU/day) and low-molecular weight (40–60 mg/day) heparin.

**Table 5:**
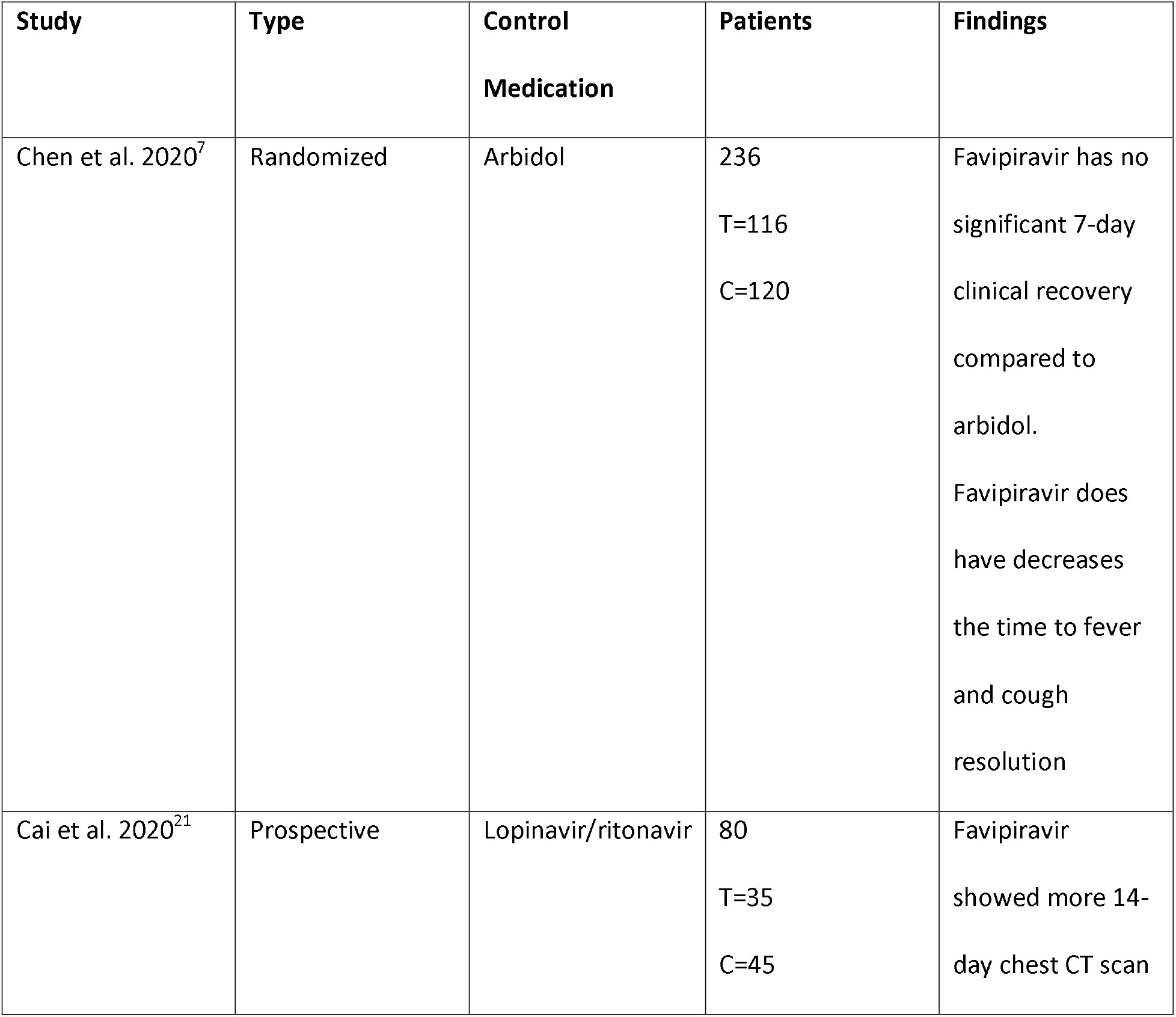
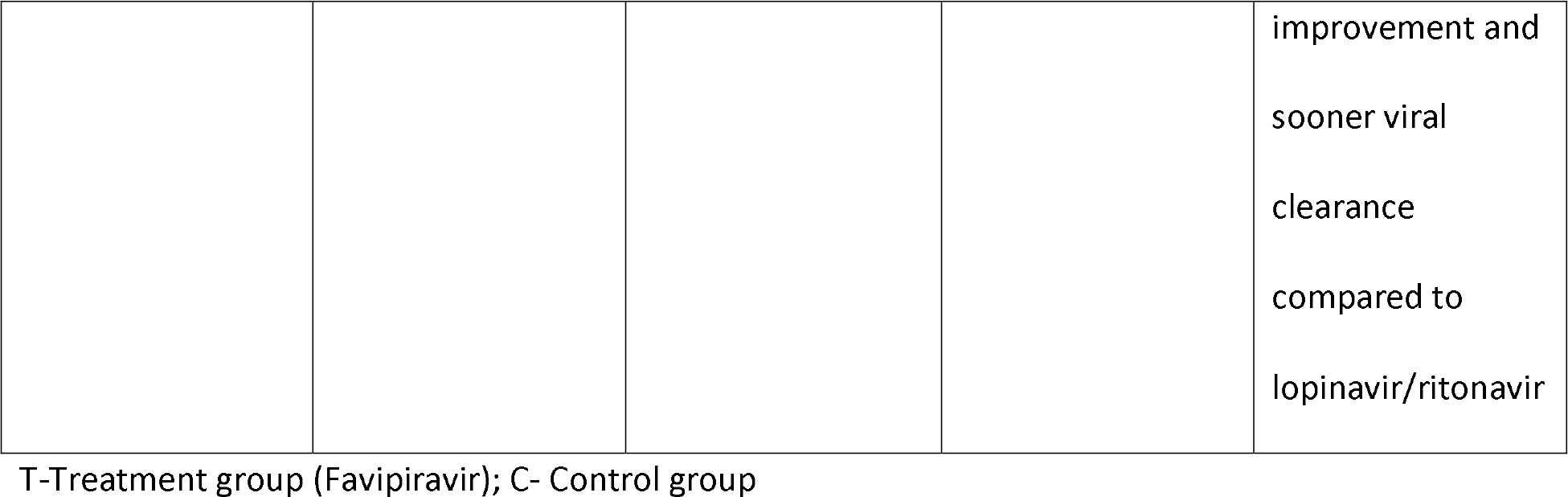
Characteristics of Favipiravir Studies for COVID-19.

| Study | Type | Control Medication | Patients | Findings |
| --- | --- | --- | --- | --- |
| Chen et al. 2020<sup>7</sup> | Randomized | Arbidol | 236<br>T=116<br>C=120 | Favipiravir has no significant 7-day clinical recovery compared to arbidol.<br>Favipiravir does have decreases the time to fever and cough resolution |
| Cai et al. 2020<sup>21</sup> | Prospective | Lopinavir/ritonavir | 80<br>T=35<br>C=45 | Favipiravir showed more 14-day chest CT scan improvement and<br>sooner viral<br>clearance<br>compared to<br>lopinavir/ritonavir |

T-Treatment group (Favipiravir); C- Control group

**Table 6:**
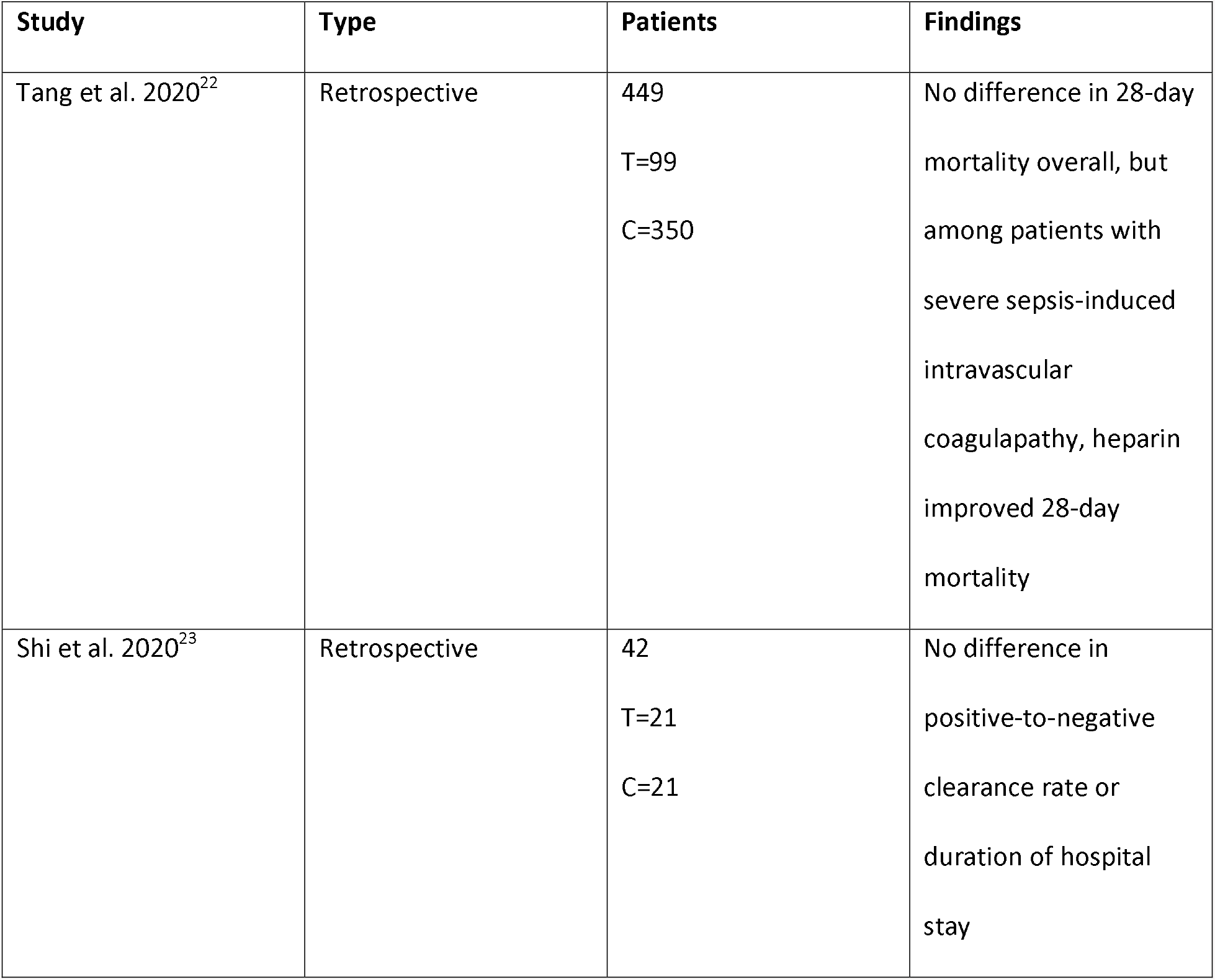
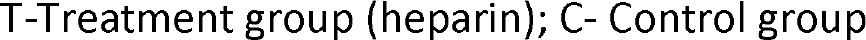
Characteristics of Heparin Studies for COVID-19.

| Study | Type | Patients | Findings |
| --- | --- | --- | --- |
| Tang et al. 2020<sup>22</sup> | Retrospective | 449<br>T=99<br>C=350 | No difference in 28-day mortality overall, but among patients with severe sepsis-induced intravascular coagulopathy, heparin improved 28-day mortality |
| Shi et al. 2020<sup>23</sup> | Retrospective | 42<br>T=21<br>C=21 | No difference in positive-to-negative clearance rate or duration of hospital stay |

#### T-Treatment group (heparin); C- Control group

Tang et al. showed no difference in 28-day mortality rates. Most patients received low-molecular weight heparin. They note significant improvement in heparin users among those with severe sepsis-induced intravascular coagulopathy. This was determined by a scoring system utilizing platelet count, prothrombin time, and Sequential Organ Failure Assessment (SOFA) scoring.[22]

Shi et al. showed no difference in outcomes including clinical improvement and positive-to-negative conversion rate. All patients in the study improved.[23]

#### Adverse Effects

The studies included in the review did not report adverse effects. However, all heparin medications have well-documented side-effects including hemorrhage, osteoporosis, renal tubular acidosis type 4 with hyperkalemia, and heparin-induced thrombocytopenia.[24–26] Adverse effects of low-molecular weight heparin are more common in patients with kidney injury.[27] Deep vein thrombosis prophylaxis presents with a lower rate of side-effects.[28]

## Discussion

Lopinavir/ritonavir, arbidol, hydroxychloroquine, favipiravir, remdesivir, and heparin are medications that have been tested in human controlled trials for COVID-19 treatment. For the meta-analyses, neither lopinavir/ritonavir nor hydroxychloroquine showed significant positive-to-negative conversion rates. The systematic review revealed inconclusive or negative results for all medications regarding

COVID-19 Treatment clinical improvement. Favipiravir showed significant improvement compared to its competitor medications, but there were no supportive therapy or placebo-controlled trials. Heparin showed significant clinical improvement only with those with severe COVID-19. Apart from heparin, the adverse effects of the medications mainly include gastrointestinal symptoms.

Lopinavir, a HIV protease inhibitor, inhibits the major protease involved in COVID-19 replication and development of functional viral proteins. Ritonavir acts to increase the levels of lopinavir and improve bioavailability.[29–31] Lopinavir/ritonavir along with ribavirin were previously used to treat SARS in nonrandomized clinical trials to prevent development of ARDS.[32] In vitro studies show an antiviral effect of lopinavir on COVID-19.[33] However, human trials show no significant difference in clinical improvement and viral shedding. Furthermore, a comparison trial shows inferiority to arbidol regarding viral clearance.[9]

While arbidol has displayed antiviral effects with previous coronaviruses,[34–35] the mechanism of action on COVID-19 is currently unknown. In human trials, arbidol shows no significant positive-negative conversion rate or recovery time compared to standard therapy or lopinavir/ritonavir.[4,9] The meta-analysis comparing seven and 14-day viral clearance between arbidol and lopinavir/ritonavir possibly favored arbidol significantly. Employing a random-effects model to account for large heterogeneity removed the statistical significance. It does show promise for post-exposure prophylaxis.[36]

Hydroxychloroquine, a member of the 4-aminoquinolines, works by neutralizing the acidic potential of lysosomes resulting in an inhibition of cell chemotaxis, phagocytosis, antigen presentation and interferon release.[37–38] In vitro studies have shown its anti-viral effects on COVID- 19, specifically by preventing viral infusion by altering the pH of cell membranes and impairing ACE2 receptor-mediated entry. It further disrupts viral activity inside the cell.[39] Combining all the hydroxychloroquine human trials showed no benefit with reducing COVID-19 viral shedding time. One retrospective trial suggests COVID-19 Treatment increase mortality with hydroxychloroquine use.[14] There are no human trials showing the efficacy of Hydroxychloroquine for COVID-19 prophylaxis. Side-effects include visual abnormalities, gastrointestinal issues, cardiac arrhythmias with QT interval prolongation, drug-induced psychosis, and leukopenia. It also interacts with various other medications, including heparin to increase the risk of the bleeding and lopinavir/ritonavir to further prolong the QT interval.[38,40]

Remdesivir is a prodrug that is metabolized into an analogue of adenosine triphosphate, allowing it to inhibit viral RNA polymerases.[41] In vitro studies exhibit its potential in combating SARS-CoV2.[41–42] A cohort study suggested potential benefit as compassionate use for severe COVID-19.[41] However, the randomized, double-blinded, placebo-controlled trial included in the review showed no statistical effect with remdesivir regarding clinical improvement, mortality, and viral load change.[20] Adverse effects were not significant among the groups. Limitations to the study included both study groups allowing for other therapies (i.e. glucocorticoids and lopinavir/ritonavir), although their use was not significantly different among the groups. Remdesivir was also started late in some of the study patients. The study was also considered underpowered.[20]

Favipiravir is a broad spectrum antiviral against RNA viruses. Inside infected host cells, it becomes phosphorylated into Favipiravir-RTP and inhibits viral RNA-dependent RNA polymerase.[43–44] Favipiravir also suppresses tumor necrosis factor – alpha (TNF-a) production.[45–46] The human COVID-19 trials with favipiravir are compared with two specific controls. Compared to arbidol, favipiravir reduces symptom duration.[7] Compared to lopinavir/ritonavir, favipiravir reduces viral shedding time and hastens chest CT scan improvement while having fewer side effects.[21] Favipiravir adverse effects include gastrointestinal symptoms and elevated uric acid levels.[7,21] Its safe profile has made it a preferred medical therapy for those with cardiovascular and renal disease.[47–48]

Heparin has various non-anticoagulant properties including reducing IL-6-associated inflammation.[49–51] IL-6 causes hypercoagulation.[51] Levels are significantly higher in severe COVID-19 patients.[49] Heparin binds to IL-6, reducing the interaction between IL-6, SIL-6R, and sgp130.[54] This benefit may explain the meta-analysis findings showing ARDS-associated mortality benefit with early low-molecular weight heparin initiation.[55] Heparin also binds to various viral entry proteins, including herpes simplex, zika, and SARS.[57–58] Similarly, it attaches to the S1 spike protein of COVID-19 and causes a conformational change, inhibiting viral membrane fusion with the cell wall.[59] The current studies suggest benefit mainly with severe COVID-19 cases.[22–23]

### Limitations

The meta-analysis portion of the study has some limitations. The first limitation is the small number of patients in the trials and therefore the overall analysis. Another limitation is the use of surrogate endpoints to complete the meta-analysis. This is regarding the use of CT scan resolution for viral clearance in the hydroxychloroquine analysis. Chest CT scans have significant negative predictive value, but is not directly comparable to RT-PCR.[16–19] The endpoints were not well-established among all reviewed medications, making it difficult to compare them between studies.

Regarding the systematic review, publication bias influences the information presented. Favipiravir trials on COVID-19 only involve those compared with other medications and not with a placebo or supportive therapy control arm. The heparin studies mainly involved the level of severity of COVID-19 rather than having the infection itself.

### Conclusion

Current investigated medications do not hasten viral clearance time. Clinical improvement is equivocal with lopinavir/ritonavir, arbidol, hydroxychloroquine, and remdesivir. Favipiravir shows faster viral clearance and clinical improvement compared to lopinavir/ritonavir and arbidol. Heparin shows benefit in patients with severe COVID-19 infections.

## Data Availability

All data is available for review

